# Evaluation of Six Commercial and Non-Commercial Colistin Resistance Diagnostics

**DOI:** 10.1101/2024.03.21.24304472

**Authors:** Tumisho Mmatumelo Seipei Leshaba, Masego Mmatli, Lebogang Skosana, Nontombi Marylucy Mbelle, John Osei Sekyere

## Abstract

**Background:** Resistance to colistin, a last-reserve antibiotic used for treating drug-resistant infections, is increasing globally. This study evaluated six diagnostic tests designed to detect colistin-resistant pathogens.

**Methods:** PCR and broth microdilution assays (BMD) were used to respectively characterize the molecular mechanisms and phenotypic colistin resistance of 142 Gram-negative bacterial isolates and controls. The sensitivity, specificity, positive- and negative-predictive values, major (ME) and very major errors (VME), categorical and essential agreements (EA) of ComASP Colistin, CHROMagar COL-*APSE*, Rapid NP Test, Sensititre, MicroScan, and Vitek 2 were determined with these isolates; the BMD was used as gold standard.

**Results:** The Vitek 2, Sensititre, and ComASP tests were more efficient, albeit with concerning ME and VMEs and low EAs. Sensititre was 100% specific with 0% ME and 3.61% VME; Vitek 2 had the least VME (1.25% and 0%) and a low EA (57.50%). ComASP had an EA of 75.35%. MicroScan was highly sensitive (96.55%) but less specific (87.50%), with very below-accetable EAs (48.11%). The CHROMAgar COL-*APSE* efficiently identified the species with their unique colours but was the least specific (67.80%), with the highest ME (32.20%) and high VME (7.23%). The Rapid NP test had the highest VME (7.84%), producing results within 4 hours with 92.16% sensitivity and 96.08% specificity.

**Conclusion:** Vitek 2, MicroScan, ComASP colistin, and Sensititre are good for determining colistin resistance; the latter two tests are recommendable for low-resourced laboratories. The in-house Rapid NP test has short turnaround time with high efficiency for initial resistance screening.

## Introduction

Increasing use of carbapenems to treat multidrug-resistant bacterial infections invariably led to the adoption of colistin as a last-reserve antibiotic to counter infections that are resistant to carbapenems [1–3]. Hence, bacteria that are resistant to colistin are increasingly being reported worldwide [4–6]. Resistance to colistin is mediated by several molecular mechanisms, including the mobile colistin resistance (*mcr*) gene, mutations in the PmrAB and PhoPQ two-component systems, and mutation(s) in the MgrB regulator in *Klebsiella pneumoniae* [1,2,4,7]. These molecular mechanisms, alongside efflux hyperexpression and porin downregulation, and/or Ecr transmembrane protein mediate phenotype colistin (polymyxin) resistance and heteroresistance [1,8,9].

To help clinicians easily detect and monitor colistin-resistant infections, several diagnostic tests and assays, both commercial and non-commercial, have been designed and developed [3,10–12]. Some of these tests (such as the BMD, ComASP™ Colistin, MicroScan, Sensititre and Vitek) are mainly MIC (minimum-inhibitory concentration)-based, measuring only the MICs of the isolates while others are non-MIC—based, purely providing a binary result of resistant or sensitive (such as the CHROMagar COL-*APSE*, and Rapid NP test) [10–12]. The Rapid NP test was designed to only detect colistin resistance in *Enterobacterales* and is therefore not useful for all Gram-negative bacteria [10,13]. As a follow-up to previous research [11], we used 142 Gram-negative isolates and controls to evaluate the performance of six colistin resistance diagnostic tests: ComASP Colistin, CHROMagar COL-*APSE*, Rapid NP test, Sensititre, Vitek 2, and the MicroScan. The BMD remains the gold standard for testing colistin MICs and resistance in bacteria [10]; hence it was used as the reference standard in these evaluation studies.

## METHODS

### Demographics and source of clinical specimen and isolates

The evaluation study was conducted on a collection of 134 Gram-negative clinical bacterial (GNB) isolates including *Enterobacterales* (n=103), *Acinetobacter baumannii* (n=21) and *Pseudomonas aeruginosa* (n=10) that were collected from the National Health Laboratory Services, Tshwane Academic Division. Eight control strains were also included to make up to 142: *Escherichia coli* MC1, *E. coli* MC2, *E. coli* MC3, *Salmonella* group D MC4, *Salmonella* group D MC5, *E. coli* (mcr-1) EMRC, *E. coli* ATCC 25922 (EATCC), *P. aeruginosa* ATCC 27853 (PATCC). Species identification was performed as part of routine laboratory testing using Vitek® 2 automated system (Biomerieux, France) (Table S1). Demographic data such as sex, age and sample source were retrieved from the NHLS TrakCare system (Table S1).

The National Food Institute, Technical University of Denmark provided five *mcr*-gene control strains (four *E. coli* with *mcr*-1, *mcr*-2, *mcr*-3 and *mcr*-4, as well as one *Salmonella* spp. with *mcr*-5) for this study. Antimicrobial susceptibility testing also included one *Escherichia coli* ATCC 25922 and one *P. aeruginosa* ATCC 27853.

The labels of the isolates used in this article are known to only the researchers as the original labels were de-identified to maintain the patients anonymity.

### Broth Microdilution

The reference minimum inhibitory concentration (MIC) of each isolate was determined by manual broth microdilution (BMD) according to ISO standard 20776-1 [14]. Colistin sulfate powder (Glentham Life Sciences, England) was diluted in cation- adjusted Mueller Hinton broth (CAMHB) in untreated 96-well microtiter polystyrene plates (Eppendorf, Germany). Dilutions to the MIC range 128-0.25 µg/mL were established. Colistin sensitivity results were interpreted according EUCAST breakpoints (susceptible ≤2Lmg/L; resistant >2Lmg/L). (https://www.nature.com/articles/s41598-020-63267-2).

### Detection of *mcr*-genes

All isolates were screened for the presence of *mcr*-1, *mcr*-2, *mcr*-3, *mcr*-4 and *mcr*-5 genes using PCR. The multiplex PCR method previously described by Rebelo *et al*. was used for the screening of *mcr*-genes [15]. The *mcr*-gene control strains supplied by the kind courtesy of Professor Rene S. Hendriksen of the National Food Institute, Technical University of Denmark, were used as positive controls.

### Vitek 2 system

Besides species identification, the Vitek was also used to determine the MICs of the isolates, using the manufacturer’s protocols.

### MicroScan

The MicroScan walkaway system (Beckman Coulter, South Africa) was used to determine the sensitivity of the species to colistin using the manufacturer’s protocol. Briefly, an overnight culture grown on blood agar was immediately standardized using the the prompt inoculation system provided in the reagent packaging for MicroScan AST (antimicrobial sensitivity testing) and identification (ID) testing. The suspension was placed into the N66 and processed overnight in the MicroScan machine.

### ComASP™ Colistin

ComASP™ Colistin by Liofilchem (Roseto degli Abruzzi, Italy) is a compact panel containing freeze-dried colistin which when diluted should result in two-fold dilutions ranging from 0.25-16 µg/mL [11,16]. The non-automated BMD-based assay allows for four samples to be tested on a single test. The manufacturer’s instructions were followed to perform the ComASP™ Colistin test. A 0.5 McFarland suspension of the isolates was prepared in a solution of 250 mL saline and then diluted to 1:20 in saline (Gibco, Thermo Fisher Scientific, USA) to obtain solution A. Solution A (0.4 mL) was added to a tube containing 3.6mL Mueller-Hinton broth provided by the ComASP™ Colistin to obtain solution B. One hundred microliters of solution B were dispensed into each well and the panels were incubated (Vacutec, US) at 36±2°C for 16 to 20 hours in ambient air. Results were read by visually analysing the plates for turbidity.

### ChromAgar Col-*APSE*

Fresh 24-hour culture of each isolate was dissolved in saline and adjusted to 0.5 McFarland’s standard. This suspension was spread on the ChromAgar Col- *APSE* plate and incubated for 18 - 24 hours. The manufacturer’s protocol was used throughout this test (https://www.nature.com/articles/s41598-020-63267-2).

### Sensititre™

Sensititre™ (Thermo Fisher Scientific, USA) “FROL” colistin-customised plates were used in this study. A 0.5 McFarland standard in saline was prepared for each isolate (Gibco, Thermo Fisher Scientific, USA). Ten microlitres of the 0.5 McFarland suspension was transferred into an 11 mL tube containing CAMHB with TES buffer for all non-intrinsic *Enterobacterales*, *A. baumanni* and *P. aeruginosa* isolates. However, for *Proteus* species, *Providencia* spp. and *Morganella* morganii 1 μL was transferred into the 11 mL CAMHB tube. Each Sensititre™ plate constitutes of eight rows with 12 wells, each row allows for the testing of one isolate at different concentrations of colistin. Wells 1-11 contain dehydrated colistin at concentrations of 0.12-128 µg/mL, respectively. Well 12 represents a positive control and therefore does not contain colistin. Each well was inoculated with 50 μL of the bacterial broth suspension using a manual pipette. Each plate was then covered with an adhesive seal and incubated (Vacutec, US) at ± 35°C in an aerobic environment. Results were read by visually analysing the plates for turbidity.

### In-house Rapid Polymyxin NP test

Rapid Polymyxin NP test is based on the detection of glucose metabolism related to *Enterobacterales* growth in the presence of a defined concentration of colistin [17]. The formation of acid metabolites is shown by a colour change of a pH indicator (red phenol) in less than 4 hours [17,18]. The rapid NP test was performed as described by Nordmann *et al*, 2016 [6,19–21]. To prepare the NP test solution, a 6.25g of CAMHB broth powder and 0.0125g of phenol red powder was added to 225 mL of distilled water. The pH of the solution was then adjusted to 6.7 by adding 1-N HCL in drops after which the solution was autoclaved. A 25 mL of 10% anhydrous D-glucose solution that had been sterilised by filtration was added to the autoclaved CAMHB solution. Colistin stock solution (1280 µg/mL) was diluted to 200 µg/mL working solution by adding 1 mL of the stock solution to 5.4 mL CAMHB. To make colistin-NP test solution, 100 µL of the colistin working solution (200 µg/mL) was added to 3900 µL NP test solution to get a final concentration of 5 µg/mL. The Rapid Polymyxin NP test was carried out in 96 well polystyrene plates (Eppendorf, Germany). A 150 µL of NP test solution alone without colistin was added to wells 1, 3, 5, 7, 9 and 11 of each row (A-H) of the 96-well microtiter plates. Wells 2, 4, 6, 8, 10 and 12 contained 150 µL of colistin-NP test solution (5 µg/mL). *Proteus mirabilis* isolate from our collection was used as a resistant control and *Escherichia coli* ATCC 25922 was used as a susceptible control.

A 3.5 McFarland standard was prepared for each test isolates with sterile 0.85% NaCl solution and used within 15 of preparation. Each *Enterobacterales* isolate was inoculated (50 µL) in parallel into the two wells, one without colistin and one with colistin. Wells H7-8 were inoculated 50 µL *P. mirabilis* and wells H9-10 were inoculated with 50 µL *E. coli* ATCC 25922. Wells H11-12 were inoculated with 50 µL of 0.85% NaCl solution. The microtiter plates were incubated for 3 h at 35L±2, after 3 h of incubation, the isolates were checked for change in colour every 15 minutes until 4 h of incubation. The results were read visually where a change in colour of phenol red (orange to bright yellow) indicated growth.

### Data analysis

All six antimicrobial susceptibility testing methods were compared to the ISO standard 20776-1 BMD. Colistin MIC results were interpreted using EUCAST’s breakpoints (susceptible, ≤2 µg/mL; resistant, >2 µg/mL). For each test, the false positives, false negatives, true positives, and true negatives were determined and used for downstream determination of the other performance indices (Table S1). We calculated the rates of essential agreement (EA), categorical agreement (CA), very major error (VME), and major error (ME) using already described methods [11]. Categorical agreement was defined as an agreement in the classification of susceptible or resistant between the evaluated test and the reference BMD. Essential agreement was defined as an MIC value within a 2-fold (one doubling) dilution of the BMD result. A very major error occurred when the tested method interpretation of an isolate was susceptible while the BMD interpretation was resistant for the same isolate. A major error occurred when the investigated method’s interpretation was resistant, and the BMD interpretation was susceptible for the same isolate. The sensitivity and specificity of each test were calculated as previously described [11].

GraphPad Prism 10 was used to calculate the odds ratio and Fisher’s exact test to determine the significance of each test’s results against the BMD.

## RESULTS

### Reference BMD

Reference MICs of all 142 isolates were determined by standard broth microdilution (BMD), either in this study or as standard protocol by the National Health Laboratory Services Tshwane Academic Division. One *A. baumannii,* 51 *Enterobacterales* and seven *P. aeruginosa* isolates were colistin-susceptible with BMD, including one *E. coli* ATCC 25922 and a *P. aeruginosa* ATCC 27853 (Table 1). The colistin-resistant isolates by BMD include 59 *Enterobacterales*, 20 *A. baumannii* and four *P. aeruginosa*. Apart from the five *mcr* control strains, only two isolates, BB2 and EMRC, had *mcr*-1 genes (Table 1). Isolate BB2 is an *A. baumannii* with a MIC of 32 µg/mL, while isolate EMRC is an *E. coli* with a MIC of 4 µg/mL. All eight intrinsically colistin-resistant *Enterobacterales* (*Proteus mirabilis, Serratia marcescens, Morganella morganii, Proteus vulgaris, Providencia stuartii, and Ralstonia/Burkholderia pickettii*) had MICs ≥ 128 μg/mL (Table 1).

### Commercial BMD methods

All 142 isolates were tested on ComASP™ Colistin and Sensititre™. ComASP™ Colistin and Sensititre™ both had a satisfactory categorical agreement of 96.48% and 97.18%, respectively. However, ComASP™ Colistin falsely detected one *K. pneumoniae* (1µg/mL) and one *P. aeruginosa* (1µg/mL) as colistin- susceptible isolates, resulting in 3.40% MEs. Sensititre™ accurately identified all colistin- susceptible isolates. However, it failed to identify three colistin-resistant isolates, therefore resulting in no MEs and 3.61% VMEs. Most isolates that were not detected by Sensititre™ and ComASP™ Colistin had MICs close to the breakpoints (4-8µg/mL). Only one isolate that was not detected by Sensititre™ had an MIC >8 µg/mL i.e., a *K. pneumoniae* with an MIC of 32µg/mL. ComASP™ Colistin had an overall sensitivity and specificity of 96.39% and 96.61%, respectively, whereas Sensititre™ had 96.39% and 100% (Table 2).

Notably, ComASP™ fared better with *Enterobacterales* than with all Gram-negative bacteria as its sensitivity (98.31%), specificity (98.04%), and CA (98.18%) increased while its ME (1.96%) and VMEs (1.72%) reduced (Table 2): the EA, however, increased from 69.09% in *Enterobacterales* to 75.35% in GNB. Sensititre’s *Enterobacterales* results were comparably similar to that of its GNB results (Table 2).

Vitek MIC results were obtained for 120 GNB and 94 *Enterobacterales*. Vitek had the highest sensitivity and the second or third highest specificity among GNB and *Enterobacterales*, respectively, for all the tests. It had the highest CA, the second lowest EA (after MicroScan), and one of the lowest VME among the six tests (Table 2). The Microscan was equally highly sensitive but less specific, with the lowest EA (46.11% and 46.91%) among all the MIC- based tests. Its sensitivity was 96.55% and 100% for GNB and *Enterobacterales* respectively. The MicroScan’s MEs with both GNB and *Enterobacterales* (12.50% and 12.20%) were only second to that of the CHROMAgar Col-*APSE* (Table 2) and its VMEs were 3.45% (GNB) and 0% (*Enterobacterales*).

### CHROMAgar Col-APSE

A total of 96/142 isolates grew on CHROMAgar Col-*APSE*: BMD identified 19 of them as colistin susceptible, resulting in 32.20% ME and 67.80% specificity. Most of the MEs (14/19) were due to *Klebsiella* spp. The chromogenic media detected most colistin-resistant isolates (77/83), but it also produced a high percentage of VMEs (7.23%), resulting in a sensitivity of 92.77 %. Among *Enterobacterales* alone, CHROMAgar was less sensitive (91.53%) and more specific (68.63%), with fewer MEs (31.37%) and CA (80.91%) than in GNB (Table 2).

Differentiation of the isolated bacteria by the morphological appearance of their colonies was as described by the manufacturer [22]. Metallic blue (*Klebsiella*, *Enterobacter* and *Serratia* spp.), cream white (*Acinetobacter*, *Salmonella* and *Pseudomonas* spp.), pink-red (*E. coli*) colonies were observed (Fig. 1) [22]. Furthermore, *E. coli* could be distinguished as fermenting (blue with pink borders) and non-fermenting strains (pink). Swarming was observed on the *P. mirabilis*-inoculated plate because the plate was not supplemented with p-nitrophenyl glycerol as recommended by Abdul Momin *et al* (2017) (Figure 1) [22].

**Figure 1.**
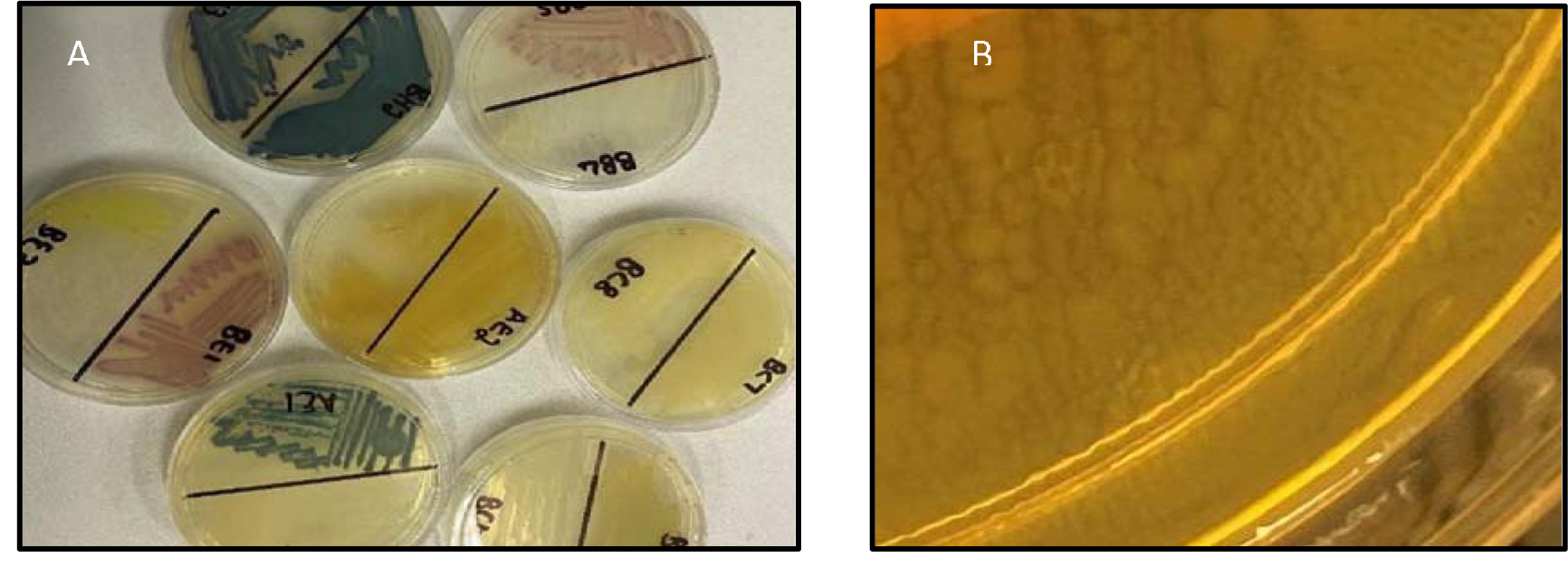
Appearance of different bacterial species on the CHROMAagar-colAPSE chromogenic media. (A) Bacterial growth on CHROMAgar Col-APSE showing different colony colours. (B) Swarming by P. mirabilis on CHROMAgar Col- APSE

### In-house Rapid NP Test

Only 102/142 of the isolates that were *Enterobacterales* were included in this test. This included 51 colistin-susceptible and 51 non-intrinsically colistin-resistant *Enterobacterales* isolates. The overall sensitivity and specificity of the in-house Rapid polymyxin NP test was 92.16% and 96.08% respectively. Colistin-resistant isolates that were not detected include one *Salmonella* spp., two *K. pneumoniae* and one *E. coli*, with MICs ranging from 4 – 128 µg/mL. Two of the false-resistant isolates, also grew on CHROMAgar Col-*APSE* (Table S1). In this study, most results were positive within two hours of incubation. Six isolates, however, were not detectable after three hours of incubation but were positive after four hours. Amongst the slow-growing isolates were four *Salmonella* spp., one *E. coli,* and one *E. cloacae*.

## DISCUSSION

The proliferation of colistin resistance among GNB has necessitated the development of better diagnostics to quickly and efficiently detect colistin-resistant organisms to control their spread [1,2,4,10,11]. This study evaluated six antimicrobial susceptibility tests that were developed to detect colistin-resistance with efficiency. Broth microdilution is currently the golden standard for colistin susceptibility testing; however, it is considered tedious and difficult [10]. Vitek 2, ComASP™ Colistin, and MicroScan had the best detection (sensitivity) of colistin-resistant isolates (Table 2). When compared with Sensititre™, CHROMagar Col-*APSE*, the Rapid Polymyxin NP test, ComASP™ Colistin, and MicroScan, Vitek 2 produced the least VMEs at 1.25% and 0% for GNB and *Enterobacterales*, respectively. This was followed by the MicroScan and ComASP, with VMEs of 3.45% vrs 3.57% and 0% vrs 1.96% respectively for all isolates and *Enterobacterales* (Table 2).

In essence, the Vitek 2, which is common in many well-resourced routine diagnostic microbiology laboratories, is very efficient in detecting colistin resistance albeit its low EA should be considered when using MICs from Vitek 2. Moreover, its initial cost, long turnaround time, and skill required to operate it make it inaccessible to low-resourced laboratories. Although the MicroScan is also highly sensitive with low VMEs (particularly among *Enterobacterales*), its lower specificity and EA, 18 – 24-hour turnaround time, operating skill required, and initial cost are also major disadvantages for low-resourced laboratories [10–12]. The ComASP and Sensititre are two affordable commercial kits that can be adopted in low-resourced laboratories for colistin resistance diagnosis. The Sensititre had 100% specificity and 72.54% vrs 71.82% EA for both GNB and *Enterobacterales*, which were the best (with ComASP) among all the six tests. Hence, in terms of cost and required skill, the Sensititre and ComASP are good alternatives for determining the colistin MICs and resistance.

The EA recorded in this study for both commercial BMD methods is below the recommended EA (≥ 90%) for antimicrobial susceptibility testing systems. The EA did not improve when analysing *Enterobacterales* alone for ComASP™ Colistin, MicroScan, and Sensititre™; Vitek 2, however, had an increase from 57.50% (Gram-negative bacteria) to 59.78% (*Enterobacterales*) (Tables 1-2). ComASP™ Colistin tended to decrease or increase the MIC of a significant number of colistin-susceptible isolates (29/59) by one two-fold dilution (Table S1). Notwithstanding, this observation did not change the CA between ComASP™ Colistin and the reference BMD.

In this study, CHROMAgar Col-*APSE* produced the highest rate of MEs and a high VME second only to that of the in-house Rapid NP test (Tables 1-2), particularly for *Klebsiella* spp. Although the specificity recorded in this study and that of Osei Sekyere *et al* (at 66,67%) agree, two other studies recorded a significantly higher specificity of ≥ 97% [11,22,23]. These discrepancies could be attributed to the different bacterial inoculum concentrations used. Ali *et al* demonstrated that when a 0.5 McFarland standard inoculum was used, colistin-susceptible isolates were able to grow on the media [23], whereas the same isolates were inhibited when the inoculum was diluted to a density of 1×105 CFU/mL. Therefore, the high rate of MEs in this present study could be due to the inoculum used being a 0.5 McFarland standard (1.5×108 CFU).

The in-house Rapid Polymyxin NP test was expected to have the best performance for *Enterobacterales*. Even though the test was designed specifically for colistin susceptibility testing on *Enterobacterales*, its performance was inferior to that of commercial BMD methods (Tables 1-2). Compared with other studies that have achieved a sensitivity of ≥ 97%, the overall performance of the in-house Rapid Polymyxin in this study (∼92%) is slightly poor [17,18,24]. The CA of all the tests evaluated in this study except for CHROMAgar Col- *APSE*, is within the recommended standard (≥ 90%) for antimicrobial susceptibility testing systems. Nevertheless, except for Vitek 2 for all GNB and MicroScan in only *Enterobacterales*, all the other evaluated tests had an unacceptable VME rate of ≥ 1.5%. Only Sensititre™ had an acceptable rate of MEs when all isolates were considered, but ComASP™ Colistin’s ME improved when only *Enterobacterales* were examined.

Therefore, for low-resourced laboratories and research institutions, the in-house Rapid NP test is a better option in terms of turnaround time, cost, and efficiency than the CHROMAgar COL-*APSE,* except that the latter has advantages of easy use and species identification through its chromogenic compounds. In terms of cost and turnaround time, the CHROMAgar is comparable to the two other commercial MIC tests: Sensititre and ComASP. Nevertheless, the higher efficiency of the former two and their ability to provide actual MICs make them recommendable. Yet, CHROMAgar is easier to use than these two MIC tests and the MICs of ComASP should be used with caution among *Enterobacterales* owing to its 75.35% EA.

The statistical odd ratio and *p-*values metrics indicate that the tests, specifically the Vitek 2, Sensititre™, ComASP™ Colistin, MicroScan, and the in-house Rapid NP test, have a relatively high level of performance in identifying both resistant and sensitive isolates among GNB. The PPV and NPV suggest that the tests’ predictive values are highly reliable. The odds ratio suggests a very strong association between the test results and the resistance being tested for, indicating that the likelihood of detecting resistant isolates is significantly higher when they are truly resistant, as compared to when they are not. The extremely low p-value indicates that this finding is statistically significant, meaning there’s a very small chance that these results could have occurred by random variation alone.

## CONCLUSION

This study discovered that commercial BMD methods had the best overall performance. In the absence of resources to purchase the Vitek 2 or the MicroScan, we recommend the ComASP™ Colistin and Sensititre™ as potential alternative MIC tests for routine colistin antimicrobial susceptibility testing in clinical laboratories. However, isolates with MICs close to the breakpoint may be misinterpreted by commercial BMD tests. As the in-house Rapid NP test had the quickest turnaround time, we recommend it for colistin susceptibility testing on *Enterobacterales*, particularly in laboratories with limited resources and labour skill. Notably, CHROMAgar Col-APSE produced the most errors; with colistin being a last-reserve antibiotic, minimal error is critical. Hence, the use of CHROMAgar Col-*APSE* should be validated further, possibly with lower inoculum. Notably, all the tests were able to detect colistin resistance in *mcr-*positive strains.

## Data Availability

All data produced in the present work are contained in the manuscript

## Acknowledgements

We are grateful to the staff of the NHLS-TAD for their support and assistance in collecting and plating the isolates. We hereby regretfully report the death of our colleague Professor Nontombi Marylucy Mbelle, who died during the course of this investigation. This work is dedicated to her memory. We are ever grateful to Professor Rene S. Hendriksen, Ana Rita Bastos Rebelo, and Troels Ronco of the National Food Institute, Technical University of Denmark, for their donation of the *mcr-1* to *mcr-5* positive controls.

## Transparency declaration

The authors declare no conflict of interest.

## Author contributions

TMSL undertook the laboratory investigations, undertook initial analysis of the data and drafted the manuscript. MM assisted with the laboratory work. BLS assisted with isolates collection and plating. NMM co-supervised the study. JOS designed and supervised the study, undertook data analysis, and wrote the manuscript. All authors reviewed the final version of the manuscript.

## Ethical approval

The study was approved by the Research Ethics Committee, Faculty of Health Sciences, University of Pretoria, under reference number 550/2020. The study complied with the ICH-GCP guidelines and the Helsinki declaration.

---

Table S1. Demographics, species, complete antibiotic resistance results data, and minimum inhibitory concentrations (MICs) of the isolates used in this study.

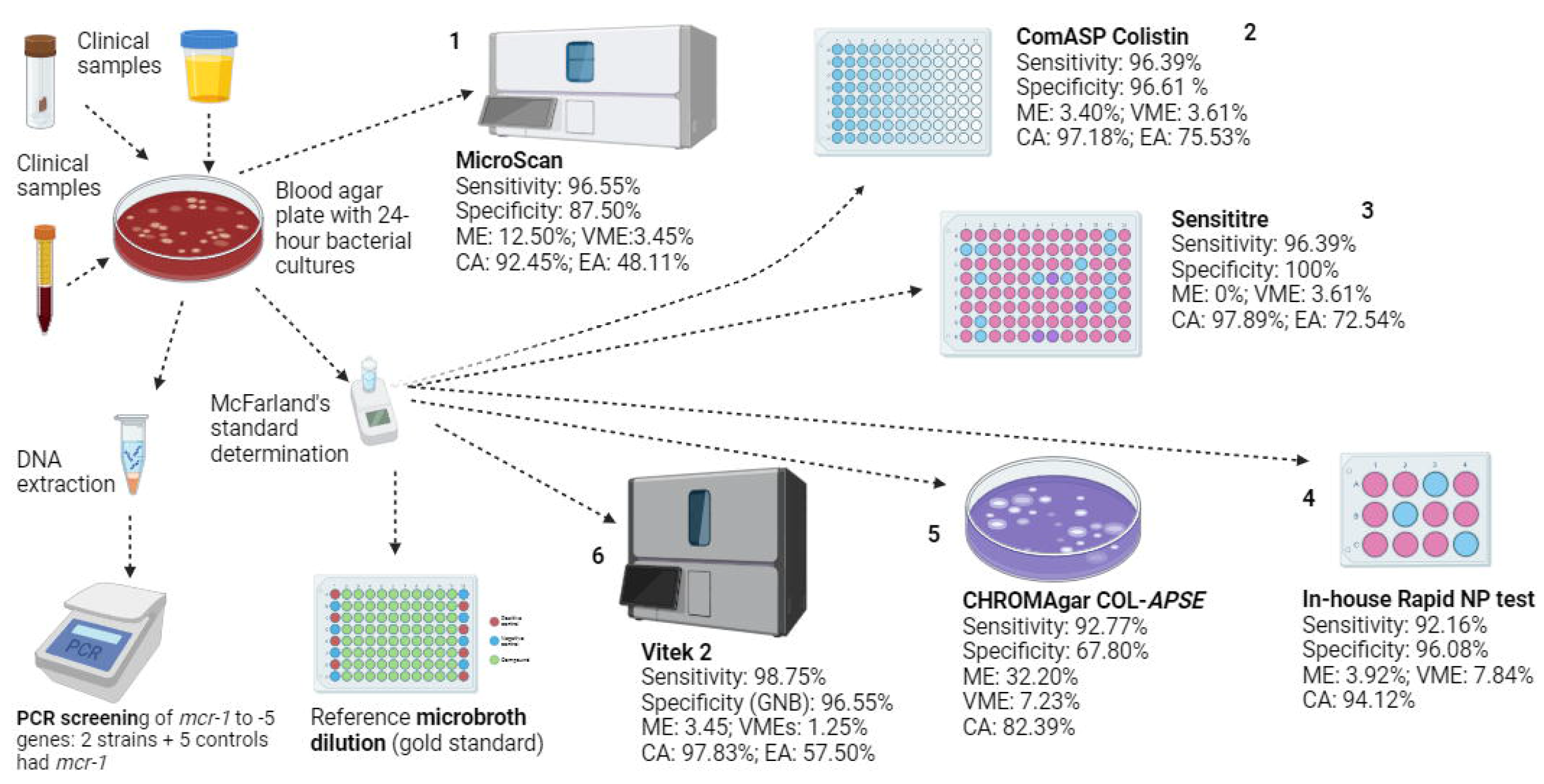

