## Supplementary material for "Evaluation of Six Commercial and Non-Commercial Colistin Resistance Diagnostics": Table 1

**Table 1. Colistin resistance detection performance results of the six tests on 142 Gram-negative bacterial isolates.**

| Isolate | **Species** | BMD MIC (µg/mL) | ComASP MIC (µg/mL) | ComASP™️ MIC Interpretation | CHROMagar™️ COL-APSE Result | NP Test Result | Sensititre MIC (µg/mL) | Sensititre MIC Interpretation | Microscan MIC (µg/mL) | Microscan MIC Interpretation | Vitek® 2 MIC (µg/mL) | Vitek® 2 MIC Interpretation | *mcr* 1-5 PCR Results |
| --- | --- | --- | --- | --- | --- | --- | --- | --- | --- | --- | --- | --- | --- |
| A1 | *E. cloacae* complex | 2 | 1 | S^[1]^ | S | S | 1 | S |  |  | 0.5 | S | NEG^[2]^ |
| A2 | *E. coli* | 0.5 | 0.5 | S | S | S | ≤ 0.25 | S | ≤2 | S | 0.5 | S | NEG |
| A3 | *S. flexneri* | ≤ 0.25 | 0.5 | S | S | S | ≤ 0.25 | S | ≤2 | S | 0.5 | S | NEG |
| A4 | *E. coli* | 0.5 | 1 | S | S | S | ≤ 0.25 | S | ≤2 | S | 0.5 | S | NEG |
| A5 | *E. coli* | 0.5 | 1 | S | S | S | ≤ 0.25 | S | ≤2 | S | 0.5 | S | NEG |
| A6 | *P. aeruginosa* | 1 | 1 | S | S |  | 1 | S | ≤2 | S | 0.5 | S | NEG |
| A7 | *P. aeruginosa* | 2 | 1 | S | R^[3]^ |  | ≤ 0.25 | S | ≤2 | S | 0.5 | S | NEG |
| A8 | *K. pneumoniae* | 1 | 4 | R | R | S | 1 | S | ≤2 | S | 0.5 | S | NEG |
| A9 | *E. coli* | 1 | 1 | S | S | S | 1 | S | ≤2 | S | 0.5 | S | NEG |
| B1 | *P. aeruginosa* | ≤ 0.25 | 0.5 | S | R |  | 0.5 | S | ≤2 | S | 0.5 | S | NEG |
| B2 | *P. aeruginosa* | 1 | 1 | S | S |  | ≤ 0.25 | S | ≤2 | S | 0.5 | S | NEG |
| B3 | *E. coli* | 0.5 | 1 | S | S | S | ≤ 0.25 | S | ≤2 | S | 0.5 | S | NEG |
| B4 | *E. cloacae* complex | 1 | 0.5 | S | S | S | ≤ 0.25 | S |  |  | 0.5 | S | NEG |
| B5 | *E. coli* | 0.5 | 1 | S | S | S | ≤ 0.25 | S | ≤2 | S | 0.5 | S | NEG |
| B6 | *E. coli* | ≤ 0.25 | 1 | S | S | S | ≤ 0.25 | S | ≤2 | S | 0.5 | S | NEG |
| B7 | *E. coli* | 1 | 1 | S | S | S | ≤ 0.25 | S | ≤2 | S | 0.5 | S | NEG |
| B8 | *E. coli* | 0.5 | 1 | S | S | S | 0.5 | S | >4 | R | 0.5 | S | NEG |
| B9 | *E. coli* | 0.5 | 1 | S | S | S | 0.5 | S | ≤2 | S | 0.5 | S | NEG |
| C1 | *K. oxytoca* | 0.5 | ≤ 0.25 | S | R | R | 0.5 | S | >4 | R | 0.5 | S | NEG |
| C2 | *K. pneumoniae* | 0.5 | 0.5 | S | R | S | ≤ 0.25 | S | ≤2 | S | 0.5 | R | NEG |
| C3 | *E. coli* | 0.5 | 0.5 | S | S | S | ≤ 0.25 | S |  |  | 0.5 | S | NEG |
| C4 | *E. coli* | ≤ 0.25 | 0.5 | S | S | S | ≤ 0.25 | S | ≤2 | S | 0.5 | S | NEG |
| C5 | *K. pneumoniae* | 0.5 | 0.5 | S | R | S | ≤ 0.25 | S | ≤2 | S | 0.5 | S | NEG |
| C6 | *K. pneumoniae* | 2 | 0.5 | S | R | S | 0.5 | S | 4 | R | 0.5 | S | NEG |
| C7 | *K. oxytoca* | 2 | 2 | S | R | S | ≤ 0.25 | S | ≤2 | S | 0.5 | S | NEG |
| C8 | *E. cloacae* complex | ≤ 0.25 | 0.5 | S | S | S | ≤ 0.25 | S |  |  | 1 | S | NEG |
| C9 | *E. cloacae* complex | ≤ 0.25 | 0.5 | S | R | S | ≤ 0.25 | S |  |  | 0.5 | S | NEG |
| D1 | *E. coli* | 1 | 0.5 | S | S | S | 0.5 | S | 4 | R | 0.5 | S | NEG |
| D2 | *K. pneumoniae* | 0.5 | 0.5 | S | R | S | 0.5 | S | ≤2 | S | 0.5 | S | NEG |
| D3 | *K. pneumoniae* | 0.5 | ≤ 0.25 | S | R | S | 0.5 | S | ≤2 | S | 0.5 | S | NEG |
| D4 | *E. cloacae* complex | 2 | 0.5 | S | R | S | 0.5 | S |  |  | 0.5 | S | NEG |
| D5 | *E. coli* | 2 | 0.5 | S | S | S | 0.5 | S | ≤2 | S | 0.5 | S | NEG |
| D6 | *E. coli* | 2 | 0.5 | S | S | S | 0.5 | S | ≤2 | S | 0.5 | S | NEG |
| D7 | *E. coli* | 1 | 0.5 | S | S | S | 0.5 | S | ≤2 | S | 0.5 | S | NEG |
| D8 | *K. oxytoca* | ≤ 0.25 | ≤ 0.25 | S | R | R | ≤ 0.25 | S | ≤2 | S | 0.5 | S | NEG |
| D9 | *E. coli* | 2 | 0.5 | S | S | S | 0.5 | S |  |  | 0.5 | S | NEG |
| E1 | *E. coli* | 0.5 | 0.5 | S | S | S | 0.5 | S | ≤2 | S | 0.5 | S | NEG |
| E2 | *E. coli* | 0.5 | 0.5 | S | S | S | 0.5 | S | 4 | R | 0.5 | S | NEG |
| E3 | *K. pneumoniae* | 0.5 | 0.5 | S | R | S | 0.5 | S |  |  | 0.5 | S | NEG |
| E4 | *E. coli* | 1 | 0.5 | S | S | S | 1 | S | ≤2 | S | 0.5 | S | NEG |
| E5 | *C. freundii* | 0.5 | 0.5 | S | S | S | ≤ 0.25 | S | ≤2 | S | 0.5 | S | NEG |
| E6 | *E. cloacae* complex | 2 | ≤ 0.25 | S | S | S | ≤ 0.25 | S |  |  | 0.5 | S | NEG |
| E7 | *E. coli* | 2 | 0.5 | S | S | S | 1 | S | ≤2 | S | 0.5 | S | NEG |
| E8 | *E. cloacae* complex | 1 | 0.5 | S | S | S | 1 | S | ≤2 | S | 0.5 | S | NEG |
| E9 | *C. koseri* | 2 | 1 | S | S | S | 1 | S | ≤2 | S | 0.5 | S | NEG |
| F1 | *E. cloacae* complex | 1 | ≤ 0.25 | S | S | S | 1 | S | ≤2 | S | 0.5 | S | NEG |
| F2 | *E. coli* | 2 | 0.5 | S | S | S | 2 | S | ≤2 | S | 0.5 | S | NEG |
| F3 | *E. coli* | 1 | 0.5 | S | S | S | 1 | S | ≤2 | S | 0.5 | S | NEG |
| F4 | *K. pneumoniae* | 1 | 0.5 | S | R | S | 1 | S | ≤2 | S | 0.5 | S | NEG |
| F5 | *K. pneumoniae* | 1 | 0.5 | S | R | S | 1 | S | ≤2 | S | 0.5 | S | NEG |
| F6 | *K. pneumoniae* | 1 | 0.5 | S | R | S | 1 | S | ≤2 | S | 0.5 | S | NEG |
| F7 | *K. pneumoniae* | 1 | 2 | S | R | S | 1 | S | ≤2 | S | 0.5 | S | NEG |
| F8 | *P. aeruginosa* | 1 | 0.5 | S | R |  | 1 | S | ≤2 | S | 0.5 | S | NEG |
| F9 | *P. aeruginosa* | 1 | 8 | R | S |  | 1 | S | 4 | R | 1 | S | NEG |
| G1 | *E. cloacae* complex | 1 | 0.5 | S | S | S | ≤ 0.25 | S |  |  |  | S | NEG |
| G4 | *E. coli* | 1 | 1 | S | S | S | 0.5 | S | ≤2 | S | 0.5 | S | NEG |
| BA1 | *A. baumannii* complex | 64 | >16 | R | R |  | 32 | R | >4 | R | >=16 | R | NEG |
| BA2 | *A. baumannii* complex | 64 | >16 | R | R |  | 64 | R |  |  | >=16 | R | NEG |
| BA3 | *A. baumannii* complex | 32 | >16 | R | R |  | 32 | R | 4 | R | >=16 | R | NEG |
| BA4 | *E. cloacae* complex | 64 | >16 | R | S | R | 64 | R | >4 | R | 4 | R | NEG |
| BA5 | *K. pneumoniae* | 128 | >16 | R | R | R | 128 | R | >4 | R | >=16 | R | NEG |
| BA6 | *K. pneumoniae* | 16 | 16 | R | R | R | 16 | R | >4 | R | >=16 | R | NEG |
| BA7 | *K. pneumoniae* | 32 | >16 | R | R | S | 2 | S | >4 | R | 4 | R | NEG |
| BA8 | *Salmonella* group D | 8 | 16 | R | R | R | 2 | S |  |  | 8 | R | NEG |
| BA9 | *A. baumannii* complex | 64 | 4 | R | R |  | 32 | R |  |  |  | S | NEG |
| BB1 | *E. coli* | >128 | >16 | R | R | R | >128 | R |  |  | >=16 | R | NEG |
| BB2 | *A. baumannii* complex | 32 | >16 | R | R |  | 128 | R | >4 | R | >=16 | R | *mcr*-1 |
| BB3 | *Salmonella* group D | 8 | 8 | R | R | R | 8 | R |  |  | 4 | R | NEG |
| BB4 | *A. baumannii* complex | 4 | 4 | R | S | R | 2 | S | ≤2 | S | 0.5 | S | NEG |
| BB5 | *E. coli* | 8 | ≤ 0.25 | S | R | R | 4 | R | >4 | R |  | R | NEG |
| BB6 | *K. pneumoniae* | 16 | >16 | R | R | S | 4 | R | >4 | R |  | R | NEG |
| BB8 | *A. baumannii* complex | 64 | >16 | R | R |  | 32 | R | >4 | R | >=16 | R | NEG |
| BB9 | *A. baumannii* complex | 8 | 16 | R | R |  | 8 | R | >4 | R | >=16 | R | NEG |
| BC1 | *Salmonella* group D | 128 | >16 | R | R | S | 4 | R |  |  |  | R | NEG |
| BC2 | *A. baumannii* complex | 16 | 16 | R | R |  | 16 | R | >4 | R | >=16 | R | NEG |
| BC3 | *P. aeruginosa* | 4 | 2 | S | R | NA | 4 | R | ≤2 | S | >=16 | R | NEG |
| BC4 | *Salmonella* group D | 4 | 4 | R | R | R | 8 | R |  |  |  | R | NEG |
| BC5 | *A. baumannii* complex | 16 | 16 | R | R |  | 16 | R | >4 | R | >=16 | R | NEG |
| BC6 | *A. baumannii* complex | 4 | >16 | R | R |  | 32 | R | >4 | R | >=16 | R | NEG |
| BC7 | *A. baumannii* complex | 32 | >16 | R | R |  | 32 | R | >4 | R |  | R | NEG |
| BC8 | *A. baumannii* complex | 32 | >16 | R | R |  | 32 | R | >4 | R | >=16 | R | NEG |
| BC9 | *Salmonella* group D | 4 | 8 | R | R | R | 8 | R |  |  |  | R | NEG |
| BD1 | *K. pneumoniae* | 64 | >16 | R | R | R | 64 | R | >4 | R | >=16 | R | NEG |
| BD2 | *Salmonella* group D | 4 | 4 | R | R | R | 4 | R |  |  | 4 | R | NEG |
| BD3 | *K. pneumoniae* | 16 | >16 | R | R | R | 16 | R | >4 | R | >=16 | R | NEG |
| BD4 | *E. coli* | 16 | 8 | R | R | S | 8 | R | >4 | R | >=16 | R | NEG |
| BD5 | *K. pneumoniae* | 16 | >16 | R | R | R | 16 | R | >4 | R | >=16 | R | NEG |
| BD6 | *K. pneumoniae* | 16 | 16 | R | R | R | 16 | R | >4 | R |  | R | NEG |
| BD7 | *K. pneumoniae* | 16 | 4 | R | R | R | 16 | R | >4 | R | >=16 | R | NEG |
| BD8 | *Salmonella* group D | 8 | 16 | R | R | R | 8 | R |  |  |  | R | NEG |
| BD9 | *K. pneumoniae* | 16 | >16 | R | R | R | 16 | R | >4 | R | >=16 | R | NEG |
| BE1 | *E. coli* | 16 | 8 | R | R | R | 8 | R | >4 | R | >=16 | R | NEG |
| BE2 | *P. aeruginosa* | 4 | 1 | S | R |  | 8 | R | >4 | R | >=16 | R | NEG |
| BE3 | *K. pneumoniae* | 64 | >16 | R | R | R | 128 | R | >4 | R |  | R | NEG |
| BE4 | *A. baumannii* complex | 0.5 | 0.5 | S | S |  | 0.5 | S |  |  | 0.5 | R | NEG |
| BE5 | *A. baumannii* complex | 64 | >16 | R | R |  | >128 | R | >4 | R | 4 | R | NEG |
| BE6 | *E. cloacae* complex | 16 | 16 | R | R | R | 16 | R |  |  | >=16 | R | NEG |
| BE7 | *E. cloacae* complex | 64 | >16 | R | S | R | 128 | R |  |  | >=16 | R | NEG |
| BE8 | *K. pneumoniae* | 64 | >16 | R | R | R | 64 | R |  |  | 4 | R | NEG |
| BE9 | *Salmonella* group D | 4 | 4 | R | R | R | 4 | R |  |  | 4 | R | NEG |
| BF1 | *P. aeruginosa* | >64 | >16 | R | R |  | 128 | R |  |  | >=16 | R | NEG |
| BF2 | *K. pneumoniae* | 16 | 16 | R | R | R | 16 | R | >4 | R | >=16 | R | NEG |
| BF3 | *E. cloacae* complex | >64 | >16 | R | R | R | 128 | R |  |  | 4 | R | NEG |
| BF4 | *A. baumannii* complex | 64 | >16 | R | R |  | 64 | R | >4 | R | >=16 | R | NEG |
| BF5 | *A. baumannii* complex | >64 | >16 | R | R |  | 128 | R | >4 | R | >=16 | R | NEG |
| BF6 | *K. pneumoniae* | 8 | 16 | R | R | R | 16 | R | >4 | R | >=16 | R | NEG |
| BF7 | *K. pneumoniae* | 64 | 8 | R | R | R | 32 | R | 4 | R | >=16 | R | NEG |
| BF8 | *K. pneumoniae* | 32 | 16 | R | R | R | 16 | R | >4 | R |  | R | NEG |
| BF9 | *K. pneumoniae* | 16 | >16 | R | R | R | 16 | R |  |  | >=16 | R | NEG |
| BG1 | *A. baumannii* complex | 16 | 16 | R | R |  | 16 | R |  |  |  | R | NEG |
| BG3 | *K. pneumoniae* | 32 | 16 | R | R | R | 32 | R | >4 | R | >=16 | R | NEG |
| BG5 | *K. pneumoniae* | 64 | >16 | R | S | R | 64 | R | >4 | R | >=16 | R | NEG |
| BG7 | *A. baumannii* complex | 32 | 16 | R | R |  | 16 | R |  |  | >=16 | R | NEG |
| BG8 | *K. pneumoniae* | 32 | >16 | R | R | R | 32 | R | >4 | R | >=16 | R | NEG |
| BG9 | *P. aeruginosa* | 64 | >16 | R | R |  | 32 | R |  |  | 4 | R | NEG |
| BH2 | *K. pneumoniae* | 16 | >16 | R | R | R | 16 | R | >4 | R |  | R | NEG |
| BH3 | *K. pneumoniae* | 128 | >16 | R | R | R | 128 | R | >4 | R | >=16 | R | NEG |
| BH4 | *K. pneumoniae* | >64 | >16 | R | R | R | 64 | R | >4 | R |  | R | NEG |
| BH5 | *K. pneumoniae* | 64 | >16 | R | S | R | 64 | R | >4 | R | >=16 | R | NEG |
| BH6 | *A. baumannii* complex | 16 | >16 | R | R |  | 16 | R | >4 | R | >=16 | R | NEG |
| BH7 | *E. cloacae* complex | 64 | >16 | R | S | R | 64 | R |  |  | >=16 | R | NEG |
| BH8 | *K. pneumoniae* | 16 | >16 | R | R | R | 16 | R | [>4](mailto:) | R | >=16 | R | NEG |
| AA1 | *P. mirabilis* | >128 | ≥16 | R | R |  | >128 | R | >4 | R | >=16 | R | NEG |
| AA3 | *S. marcescens* | >128 | ≥16 | R | R |  | >128 | R | >4 | R | >=16 | R | NEG |
| AA4 | *M. morganii* | >128 | ≥16 | R | R |  | >128 | R | >4 | R | >=16 | R | NEG |
| AA6 | *P. vulgaris* | >128 | ≥16 | R | R |  | >128 | R | >4 | R | >=16 | R | NEG |
| AD2 | *P. stuartii* | >128 | ≥16 | R | R |  | >128 | R | >4 | R | >=16 | R | NEG |
| AD7 | *R. pickettii* | >128 | ≥16 | R | R |  | >128 | R | >4 | R | >=16 | R | NEG |
| AE1 | *S. marcescens* | >128 | ≥16 | R | R |  | >128 | R | >4 | R | >=16 | R | NEG |
| AE2 | *M. morganii* | >128 | ≥16 | R | R |  | >128 | R | >4 | R | >=16 | R | NEG |
| GA3 | *A. baumannii* complex | 64 | >16 | R | R |  | 64 | R | >4 | R | >=16 | R | NEG |
| GA4 | *E. cloacae* complex | 64 | >16 | R | R | R | 64 | R | >4 | R | >=16 | R | NEG |
| GA5 | *E. cloacae* complex | 64 | >16 | R | R | R | 64 | R | >4 | R | >=16 | R | NEG |
| CG2 | *K. pneumoniae* | 128 | >16 | R | R | R | 128 | R |  |  | >=16 | R | NEG |
| MC1 | *E. coli* | 4 | 8 | R | R | R | 4 | R |  |  | >=16 | R | *mcr-1* |
| MC2 | *E. coli* | 4 | 8 | R | R | R | 8 | R | >4 | R |  | R | *mcr-2* |
| MC3 | *E. coli* | 4 | 4 | R | R | R | 4 | R | >4 | R |  | R | *mcr-3* |
| MC4 | *Salmonella* group D | 4 | 4 | R | R | R | 4 | R | 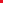 |  |  | R | *mcr-4* |
| MC5 | *Salmonella* group D | 8 | 16 | R | R | R | 8 | R |  |  |  | R | *mcr-5* |
| EMRC | *E. coli (mcr-1)* | 4 | 4 | R | R | R | 4 | R | >4 | R |  | R | *mcr-1* |
| EATCC | *E. coli* ATCC 25922 | 0.5 | 0.5 | S | S |  | 0.5 | S | 0.5 | S |  | S | NEG |
| PATCC | *P. aeruginosa* ATCC 27853 | 0.5 | 1 | S | S |  | 1 | S | 0.5 | S |  | S | NEG |

1. Sensitive
2. Negative PCR results
3. Resistant
