## Supplementary material for "Evaluation of Six Commercial and Non-Commercial Colistin Resistance Diagnostics": Table 2

***Table 2. Comparative diagnostic performance of the six tests as evaluated using Gram-negative bacteria and Enterobacterales isolates.***

| **Test** | **Sensitivity (%)** | **Specificity (%)** | **PPV^[1]^ (%)** | **NPV^[2]^ (%)** | **ME^[3]^ (%)** | **VME^[4]^ (%)** | **CA^[5]^ (%)** | **EA^[6]^ (%)** | **Odds ratio** | **Fisher’s exact test (*P-value*)** | **TAT^[7]^ (h)** |
| --- | --- | --- | --- | --- | --- | --- | --- | --- | --- | --- | --- |
| **Gram-negative bacteria** | | | | | | | | | | |  |
| **ComASP^®^ Colistin** | 96.39 | 96.61 | 97.56 | 95.00 | 3.40 | 3.61 | 97.18 | 75.35 | 756.0 | 3.20×10^−33^ | 16 - 20 |
| **CHROMagar^™^ COL-APSE** | 92.86 | 67.24 | 80.41 | 86.67 | 32.76 | 7.14 | 82.39 | N/A^[8]^ | 26.68 | 1.70×10^−14^ | 16 - 20 |
| **Sensititre** | 96.39 | 100 | 100 | 95.16 | 0 | 3.61 | 97.89 | 72.54 | Infinite^[9]^ | 1.61×10^−35^ | 16 - 20 |
| **Vitek^®^ 2** | 98.75 | 96.55 | 97.53 | 98.25 | 3.45 | 1.25 | 97.83 | 57.50 | 2212.0 | 4.49×10^−35^ | 16 - 20 |
| **MicroScan** | 96.55 | 87.50 | 90.32 | 95.45 | 12.50 | 3.45 | 92.45 | 48.11 | 196.0 | 1.49×10^−20^ | 16 - 20 |
| **Enterobacterales** | | | | | | | | | | |  |
| **ComASP^®^ Colistin** | 98.31 | 98.04 | 98.31 | 98.04 | 1.96 | 1.72 | 98.18 | 69.09 | 2950.0 | 2.24e^-29^ | 16 - 20 |
| **CHROMagar^™^ COL-APSE** | 91.53 | 68.63 | 77.14 | 87.5 | 31.37 | 8.47 | 80.91 | N/A | 24.0625 | 2.33e^-11^ | 16 - 20 |
| **Rapid NP test** | 92.16 | 96.08 | 96.00 | 92.31 | 4.00 | 7.69 | 94.12 | N/A | 288.0 | 8.53e-22 | 2 - 4 |
| **Sensititre** | 96.61 | 100.00 | 100.00 | 96.24  • | 0.00 | 3.39 | 98.18 | 71.82 | Infinity | 1.82e^-28^ | 16 - 20 |
| **Vitek^®^ 2** | 100 | 98.04 | 98.28 | 100 | 1.96 | 0.00 | 99.07 | 59.78 | Infinity | 2.75×10^−30^ | 16 - 20 |
| **MicroScan** | 100 | 87.80 | 88.37 | 100 | 12.20 | 0.00 | 93.67 | 46.91 | Infinity | 2.75×10^−17^ | 16 - 20 |

1. Positive predictive value
2. Negative predictive value
3. Major error
4. Very major error
5. Categorical agreement
6. Essential agreement
7. Turnaround time
8. Not applicable (only used for MIC tests) or results are not available owing to missing MIC data.
9. Because of a specificity of 100% and no observed false positives
